# SARS-CoV-2 infections in infants in Haiti 2020-2021; evidence from a serological cohort

**DOI:** 10.1101/2022.03.17.22272561

**Authors:** Rigan Louis, Ruiyu Pu, Tracey D Logan, Luke Trimmer-Smith, Richard Chamblain, Adriana Gallagher, Valery Madsen Beau De Rochars, Eric Nelson, Derek AT Cummings, Maureen T Long, J. Glenn Morris

## Abstract

Between February 2019 and March 2021, 388 dried blood spot samples were obtained from 257 children <30 months of age who were part of a longitudinal maternal/infant cohort in Haiti. Among the children followed, 16.7% became seropositive; sampling date was the only covariate associated with the hazard of seroconversion.

---

While there have been a number of population-based serologic studies that have assessed rates of SARS-CoV-2 infection, work to date has largely been among adults in the developed world.^1,2^ Of the 968 studies included in a recent meta-analysis, the majority (77%) of papers retained for analysis were from high income countries, which tended to have lower seroprevalence than low- to middle-income countries (LMIC).^1^ Most of the papers that have been published, irrespective of income,^3,4^ have focused on school-age or adult populations, with very limited data available on infections among the youngest age groups. However, it has also been observed that young infants have SARS-CoV-2 viral loads higher than those seen in other age groups, suggesting that they can play a critical role in transmission within a household setting.^5^ Given these circumstances, it becomes extremely important to understand the rate of infection and immune responses amongst infants in LMIC to facilitate development of appropriate surveillance and intervention strategies.

The first case of COVID-19 in Haiti was identified on March 19, 2020. As part of an international multi-site study to assess the impact of viral infection on pregnancy outcome (the ZIKAction project),^6^ our group monitored a maternal and child cohort in the Gressier region of Haiti, with dried blood spot (DBS) samples collected from infants between birth and 30 months of age between February 2019 and March 2021. Serendipitously, this time period included the first reported Haitian COVID-19 cases and the peaks of the first and second Haitian COVID-19 “waves” (Figure 1). We used these data and samples to assess serological responses to SARS-CoV-2 among study infants during the first year of the pandemic in Haiti. The predominant SARS-CoV-2 lineage circulating in Haiti during the study was B.1, with smaller numbers of cases in the B1.478 and B1.1 lineages.^7^

**Figure 1.**
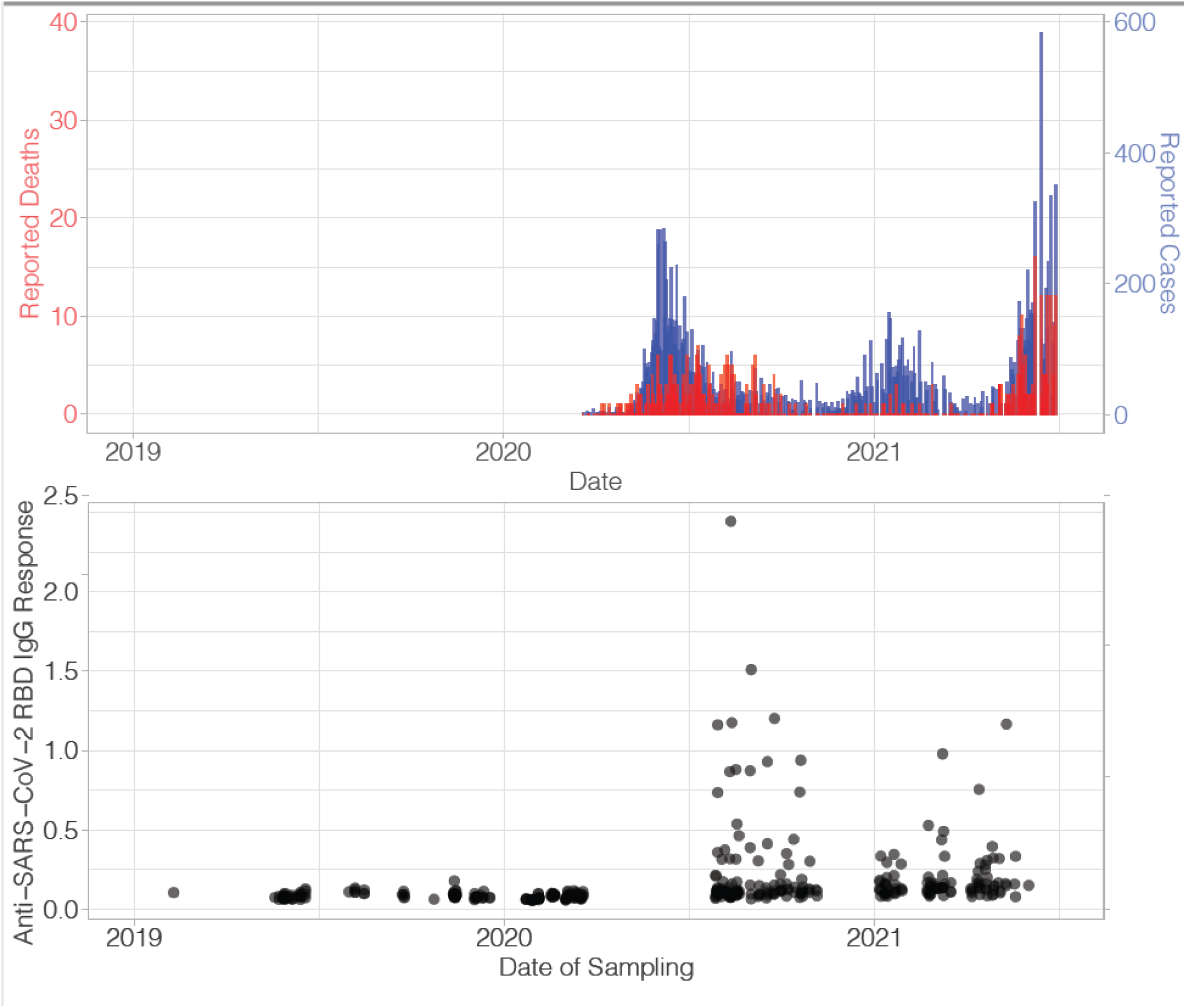
Reported cases (blue) and deaths (red) due to SARS-CoV-2 in Haiti (top) and optical density from ELISA that measures the antibody to SARS-CoV-2 Receptor Binding Domain IgG (bottom) by date of sampling for Haitian infants born between March 2019 and May 2021 and followed for up to 30 mos.

## Methods

This study was conducted under review of the University of Florida (UF) IRB and the Haitian Comite National de Bioethique of the Ministere De La Sante Publique Et De La Population. Mothers in the study were recruited at the time of their first antenatal visit; basic demographic, epidemiologic, and clinical data were obtained; and efforts were made to follow all infants born to enrolled mothers, with visits scheduled at 0, 4, 12, and 18-30 months of age. During visits, infants were screened for developmental delays, and DBS samples were collected by heel-stick. Mothers were also asked to report illness in their infants since the infant was last seen by study staff. Travel within Haiti was severely limited during the study period because of substantial political unrest and the COVID epidemic. Many mothers were also hesitant about allowing a heel-stick for blood collection. Because of this, it was not possible to obtain samples from all infants at all scheduled time points.

Detailed laboratory methods are provided in Supplemental Materials. In brief, a single 6mm biopsy punch was used to capture the area covered with each blood spot. Because the blood spots varied in size and intensity, the amount of total protein in each sample was determined by using a Bradford protein microassay. A research ELISA, adapted from a previously published protocol,^8^ was developed and used to target the SARS-CoV-2 Receptor Binding Domain (RBD).

We estimated that with 50 samples collected before the pandemic and 50 after, we had 80% power to detect a difference in seroprevalence assuming 15% of babies would be seropositive after the beginning of the pandemic and 0% before (with 95% confidence). We fit regression models to average ODs to determine factors associated with larger OD values. A mixture model was fit to the average OD’s across all samples, assuming that observations came from two distributions, those that had not been infected and those that had in the past. A threshold in OD value was identified from the mixture model that indicated a 95% probability of belonging to the distribution associated with higher values (OD>0.21). This threshold was used to define individual measurements as indicating seropositivity. Interval censored survival analysis was used to investigate the association of the hazard of SARS-CoV-2 seropositivity possible risk factors.

## Results

We obtained 388 samples from 257 children; numbers of samples by year and time point are shown in Supplemental Table 1. Longitudinal samples were obtained from 107 of these children consisting of two or more samples: this includes 84 infants for whom two samples were available, 22 with three samples, and one for whom samples were available for all four time points. The average protein concentration for all samples was 5,194 ug/mL with 18 measuring below 100 ug/mL and the highest measuring 13,209 ug/mL. There were no differences in mean protein levels when analyzed for age or year.

Using the dataset of all samples (388), a spline investigating the relationship between time of sampling and OD indicated that average ODs increased ∼0.200 in value at 400 days past January 1, 2019 or approximately March 2020 (Supplemental Figure 1). Increases in ODs that occurred in 2020 and 2021 reflected the timing of waves of reported for SARS-CoV-2 cases and deaths in Haiti over the same time periods (Figure 1). No anti-SARS-CoV-2 IgG responses above the predicted cut-off occurred before July, 2020 and no infant was seropositive at birth. Forty-three (16.7% [95% CI 12.7%-21.8%]) of 257 unique children tested seropositive at some point during the study.

We conducted a survival analysis to account for differing amounts of person time for each individual and over time (Figure 2, Supplemental Figure 2). We fit piecewise constant hazard survival models to estimate the hazard of anti-SARS-CoV-2 seropositivity over time in our study population. Hazards estimated for intervals before and after March 1, 2020 found that hazards were significantly larger in the later interval (Hazard ratio of 39.1 95% CI 13.1, 2.8e7). Age of child, mother’s age, educational attainment, marital status, urban/rural status and parity of birth were not associated with the hazard of infection. Trends in OD were similar across age groups (Supplemental Figure 3).

**Figure 2.**
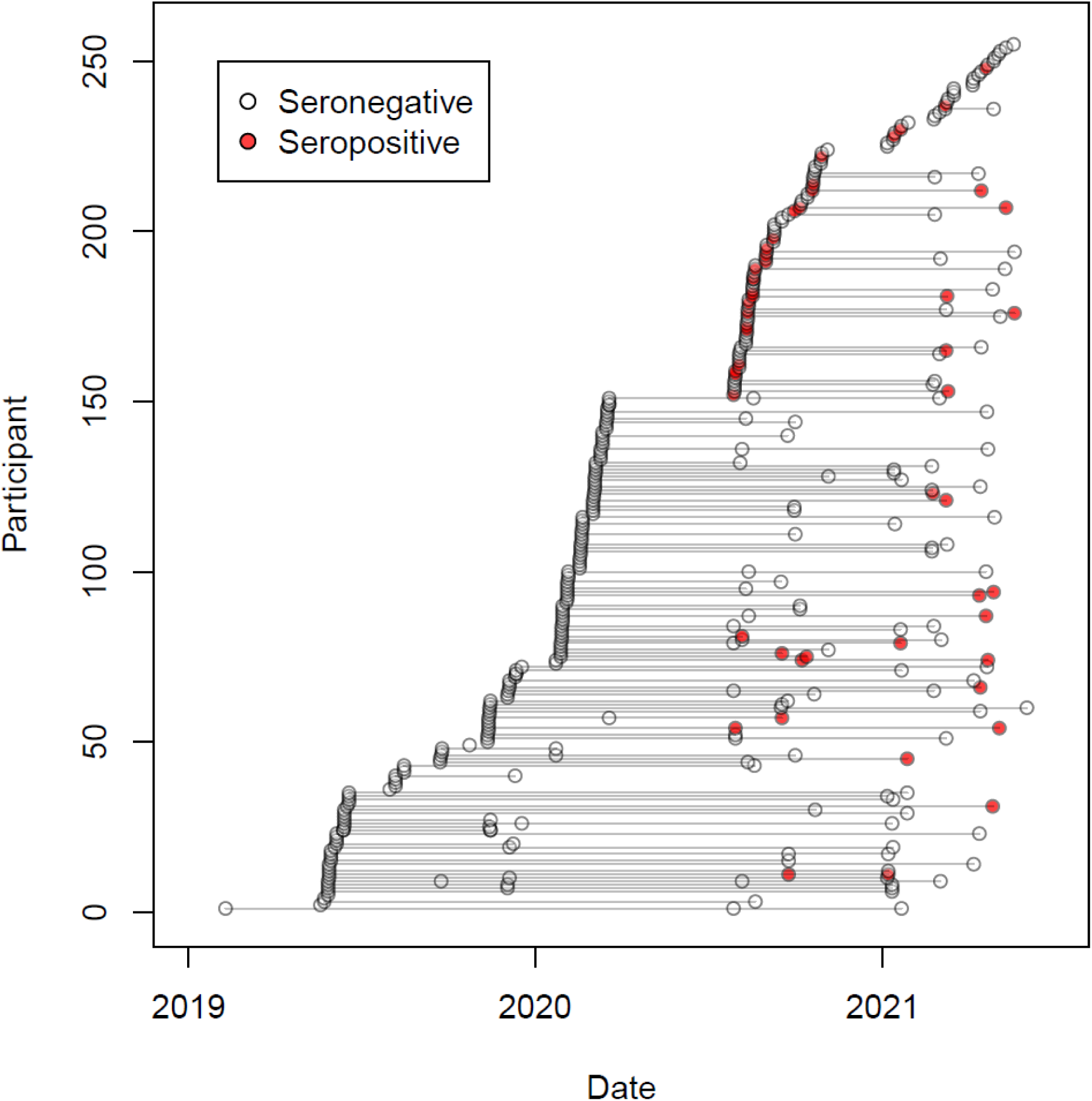
Times of follow up for each infant with serostatus indicated from 2019 to 2021. Seropositive samples are shown in red and the seronegative are without fill. Lines connect samples from the same infant.

Twenty-one of the consecutively tested children tested above the cut-off in 2020 and 2021, with seven testing above the cut-off on two of the samples. One child (of 8 total) who had a sample collected after an initial seropositive sample became seronegative upon follow up. Only limited data were available on clinical illnesses experienced by children during the study. However, no child was diagnosed at a medical facility as having COVID-19, based either on clinical presentation or laboratory testing, and mothers did not spontaneously report occurrence of more severe illnesses in their infants that might have been consistent with COVID-19.

## Comment

The overall seropositivity rate of 16.7% among our infant cohort was lower than the 39% seroprevalence previously reported by our group for adult populations in Haiti during a comparable time period.^7^ This may be a reflection of lower exposure of neonates and very young children to the general population. The adult data were collected from predominantly urban populations in Port-au-Prince. Given that studies in Peru and South Africa have demonstrated higher seroprevalence rates in more densely populated urban areas as compared with rural areas,^4,5^ caution should be used in directly comparing the data from our infants (the majority of whom came from a rural origin) with these adult data.

In work performed in middle and upper income countries, SARS-CoV-2 seropositivity in children generally identified as below the age of five has been highly variable, from less than 1% in German and Switzerland to 6% in Spain after during the first surge of the pandemic.^9-11^ Several U.S. studies examined antibody prevalence in residual blood samples of children during the late spring and early fall of 2020, with seroprevalence rates of 9.5 - 16.3%.^12,13^ Recently, a cross sectional investigation of SARS in Virginia reported 8.5% seropositivity in a study of 1,038 children; seroprevalence was highest (13.7%) in children in the 0-5 year age group.^14^ Our data are consistent with the observation that rates of seropositivity are higher in LMIC settings and underscore the widespread distribution of infection in very young children in these areas, with no risk factors identified other than occurrence in the midst of an epidemic wave.

In keeping with data showing little evidence of serious clinical illness in infected infants, we did not obtain a history of a serious preceding illness among our seropositive infants, nor were any of the children in the study formally diagnosed as having COVID-19. However, our data document that infections do occur in infants and elicit an immune response. These studies were conducted at a time when B.1 lineage strains were predominant in Haiti and need to be repeated with the successive waves of Gamma, Delta, and Omicron strains which have occurred;^7^ the health impact of these later lineages also remains to be determined. However, given that infants have been shown to have high SARS-CoV-2 viral loads,^5^ our findings underscore the potential importance of very young children in facilitating virus transmission within LMIC households and communities, and the need to consider these youngest members of society in developing models and prevention strategies for COVID-19.

## Data Availability

All data produced in the present work are contained in the manuscript

## Contributors

Conceptualization (RL, VMBDR, DATC, MTL, JGM); investigation (RL, RC, VMBDR); data curation and analysis (RL, RC, DATC, MTL, JGM); methodology (RP, TDL, LT-S, AG, EN, MTL); writing and reviewing and editing (RL, VMBDR, EN, DATC, MTL, JGM)

## Data Sharing

Contingent on adherence to IRB requirements, the complete de-identified participant dataset will be available on request. Data will be available with publication. Additional documents (study protocol, statistical analysis plan, informed consent form) will also available if needed with publication.

## Acknowledgements

This project has received funding from the European Union’s Horizon 2020 research and innovation programme under grant agreement No 734857 through the Penta Foundation (the ZIKAction Project); and by University of Florida Emerging Pathogens Institute, the Clinical and Translational Research Institute, and the Fern Audette Endowment, College of Veterinary Medicine, Gainesville FL. We thank and are grateful the provision of reagents and protocols for the rbdELISA from Aaron Schmidt, Ragon Institute/Harvard MedicalSchool, Boston MA and Jason Harris at Massachusetts General Hospital/Harvard Medical School.

---

Supplemental Figure 1: Density of observed OD values with distributions identified by a mixture model that assumed the values were drawn from two distributions, one that we associated with negative response (red) and positive response (green).

Supplemental Figure 2: Kaplan Meier survival plot showing estimated probabilities of individuals in our cohort remaining seronegative

Supplemental Figure 3: Times of follow up for each infant with serostatus indicated from 2019 to 2021. Seropositive samples are shown in red and the seronegative are without fill. Shape of points indicate age of participant (circles, at birth, squares ∼4 months, diamonds ∼12 months, triangles ∼24 months). Lines connect samples from the same infant.

Supplemental Table 1. Age of infants and year of sampling of children tested for antibodies to SARS-CoV-2 sampled in the Gessier Region between June 2019 and March 2021.

## References

1. Bobrovitz N, Arora RK, Cao C, et al. Global seroprevalence of SARS-CoV-2 antibodies: A systematic review and meta-analysis. PLoS One 2021; 16(6): e0252617.

2. Bloomfield M, Pospisilova I, Cabelova T, et al. Searching for COVID-19 Antibodies in Czech Children-A Needle in the Haystack. Front Pediatr 2020; 8: 597736.

3. Reyes-Vega MF, Soto-Cabezas MG, Cardenas F, et al. SARS-CoV-2 prevalence associated to low socioeconomic status and overcrowding in an LMIC megacity: A population-based seroepidemiological survey in Lima, Peru. EClinicalMedicine 2021; 34: 100801.

4. Kleynhans J, Tempia S, Wolter N, et al. SARS-CoV-2 Seroprevalence in a Rural and Urban Household Cohort during First and Second Waves of Infections, South Africa, July 2020-March 2021. Emerg Infect Dis 2021; 27(12): 3020–9.

5. Ochoa V, Díaz FE, Ramirez E, Fentini MC, Carobene M, Geffner J, Arruvito L, Remes Lenicov F; INBIRS COVID-19 Study Group. Infants Younger Than 6 Months Infected With SARS-CoV-2 Show the Highest Respiratory Viral Loads. J Infect Dis. 2022 Feb 1;225(3):392–395. doi: 10.1093/infdis/jiab577. PMID: 34850028; PMCID: PMC8690165.

6. Ades AE, Brickley EB, Alexander N, et al, including EC Zika Consortia Vertical Transmission Study Group. Zika virus infection in pregnancy: a protocol for the joint analysis of the prospective cohort studies of the ZIKAlliance, ZikaPLAN and ZIKAction consortia. BMJ Open 2020;10:e035307. doi: 10.1136/bmjopen-2019-035307

7. Tagliamonte MS, Mavian C, Zainabadi K, et al. Rapid emergence and spread of SARS-CoV-2 gamma (P.1) variant in Haiti. Clin Infect Dis 2021 Sep 2:ciab736. doi: 10.1093/cid/ciab736. Online ahead of print.

8. Roy V, Fischinger S, Atyeo C, et al. SARS-CoV-2-specific ELISA development. J Immunol Methods 2020; 484-485: 112832.

9. Pollán M, Pérez-Gómez B, Pastor-Barriuso R, et al. SARS-CoV-2 seroprevalence in Spain - Authors’ reply. Lancet 2020; 396(10261): 1484–5.

10. Stringhini S, Wisniak A, Piumatti G, et al. Seroprevalence of anti-SARS-CoV-2 IgG antibodies in Geneva, Switzerland (SEROCoV-POP): a population-based study. Lancet 2020. 396:313–319. doi: 10.1016/S0140-6736(20)31304-0.

11. Tönshoff B, Müller B, Elling R, et al. Prevalence of SARS-CoV-2 Infection in Children and Their Parents in Southwest Germany. JAMA Pediatr 2021; 175(6): 586–93.

12. Hobbs CV, Drobeniuc J, Kittle T, et al. Estimated SARS-CoV-2 Seroprevalence Among Persons Aged <18 Years - Mississippi, May-September 2020. MMWR Morb Mortal Wkly Rep 2021; 70(9): 312–5.

13. Bahar B, Simpson JN, Biddle C, et al. Estimated SARS-CoV-2 Seroprevalence in Healthy Children and Those with Chronic Illnesses in the Washington Metropolitan Area as of October 2020. Pediatr Infect Dis J 2021; 40(7): e272–e4.

14. Levorson RE, Christian E, Hunter B, et al. A cross-sectional investigation of SARS-CoV-2 seroprevalence and associated risk factors in children and adolescents in the United States. PLoS One 2021; 16(11): e0259823.

