## Supplemental Materials for "SARS-CoV-2 infections in infants in Haiti 2020-2021; evidence from a serological cohort"

**SUPPLEMENTAL METHODS**

**Elution of DBS Samples:** From the DBS card, a single 6 mm biopsy punch was used to capture the area covered with each blood spot. Each punch was added to a 1·0 ml microcentrifuge tube and submerged in 100 ul PBS-0·05% Tween per punch and incubated with rocking at 50 rpm at 4^o^C overnight. After incubation, the tubes were centrifuged at 10,500 X g for 2 minutes and after incubation the supernatant removed from the paper, transferred in a new microcentrifuge tube and frozen at -80^o^C until use.

**Protein quantification:** Because the blood spots varied on each card in terms of size and intensity, we determined the amount of total protein in each sample. The protein concentration was measured using a Bradford protein microassay in all DBS samples obtained in 2019 and 2020. A protein standard set consisting of BSA diluted to a working range of 0·25 to 2·0 ug/ml were created. Three two-fold dilutions of each sample were made by diluting each in PBS starting at 1:5. A commercial Bradford reagent (Maker) was added as a 1:1 dilution to each sample and incubated for 5 minutes. Absorbance was measured at 595 nm. The final protein concentration was determined by creating a standard curve plotting the 595 nm value of each BSA dilution by its corresponding concentration. The unknown concentration of each sample was calculated from the curve as well as multiplying by the dilution factor.

**SARS-CoV-2 testing:** A research ELISA targeting the Receptor Binding Domain (RBD) of the virus was used to detect exposure to SARS-CoV-2. The ELISA was adapted from a previously published protocol, which targets the RBD of the spike protein.^9^ Briefly, 96-well ELISA plates were coated with 1 ug/ml RBD protein diluted in carbonate/bicarbonate buffer (pH 9·6) and incubated at room temperature (RT) for 1 hour. Each plate was then blocked with 1X Tris-buffered saline (TBS) with 5% milk and incubated for 2 hours. After blocking, each sample, diluted at a concentration of 1:100 in TBS-0·5% Tween, was added in duplicate to the plate. Mouse anti-human IgG-HRP (Jackson Immunoresearch, 109-035-098) was then added and incubated for 1 hour. After incubation and washing, 3,3’,5,5’tretramethylbensidine (TMB, Neogen Life Sciences) was added to each well, incubated for 5 minutes then stopped using NaSO_4_. The reaction was read using a microplate reader (Multiskan FC, Fisher or SynergyH1 BioTek) for absorbance at 450nm. Included on each plate were positive controls consisting of a serum from a pool of subjects that were clinically ill with COVID and tested by rtPCR positive for the virus at least 4 weeks before collection of serum as well as a human anti-SARS-CoV-2 monoclonal antibody. A negative control consisted of a pool of serum from patients from a pre-pandemic period. Two blank wells were also included in each plate and consisted of all reagents except for primary antibody.

**Statistical analysis:** We estimated that with 50 samples collected before the pandemic and 50 after, we had 80% power to detect a difference in seroprevalence assuming 15% of babies would be seropositive after the beginning of the pandemic and 0% before (with 95% confidence). The average of ODs across the two duplicates run for each sample was calculated. We fit regression models to average ODs to determine factors associated with larger OD values including date of sample collection (represented as either year of collection or a spline term on date), age of child at sample collection, age of mother, parity of child, educational status of mother and marital status. A mixture model was fit to the average OD’s across all samples, assuming that observations came from two distributions, those that had not been infected and those that had in the past. A threshold in OD value was identified from the mixture model that indicated a 95% probability of belonging to the distribution associated with higher values (OD>0·21). This threshold was used to define individual measurements as indicating seropositivity. Interval censored survival analysis was used to investigate the association of the hazard of SARS-CoV-2 seropositivity with date of sample collection, age of child at sample collection, age of mother, parity of child, educational status of mother**.**

**Supplemental Table 1. Age of infants and year of sampling of children tested for antibodies to SARS-CoV-2 sampled in the Gessier Region between June 2019 and March 2021**.

| Year | Birth | 4-8 months | 8-18 months | 18-30 months | Total |
| --- | --- | --- | --- | --- | --- |
| 2019 | 14 | 54 | 17 | 0 | 85 |
| 2020 | 9 | 66 | 117 | 7 | 199 |
| 2021 | 0 | 0 | 56 | 48 | 104 |
| Total | 23 | 120 | 190 | 55 | 388 |

^1^Infants were enrolled to be sampled at birth, 4 months, 12 months and 24 months, however sampling variation occurred due to local political, climatic, and geological events in Haiti over the course of the study.
