## Supplementary figures and images for "SARS-CoV-2 infections in infants in Haiti 2020-2021; evidence from a serological cohort"

### Figure S1

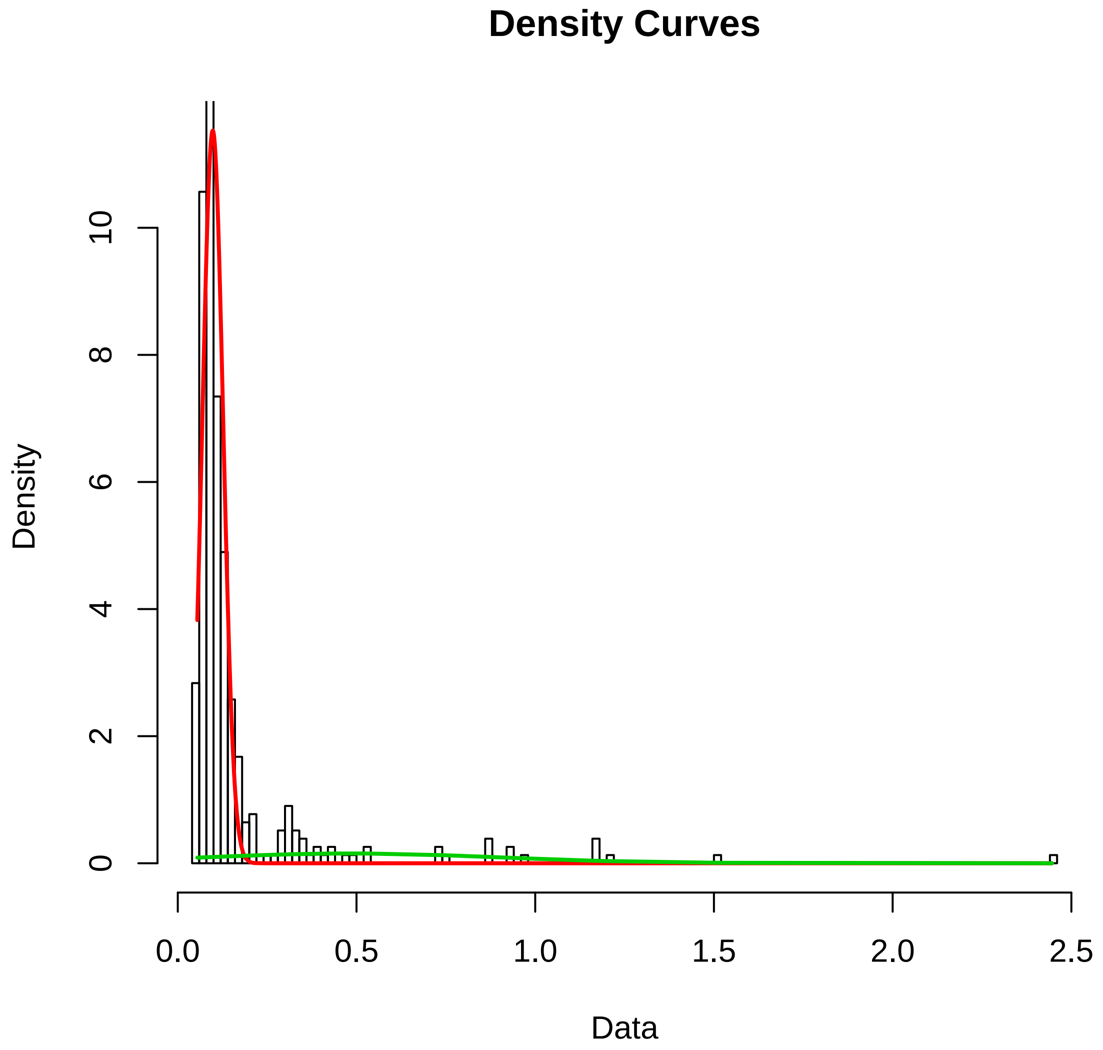

### Figure S2

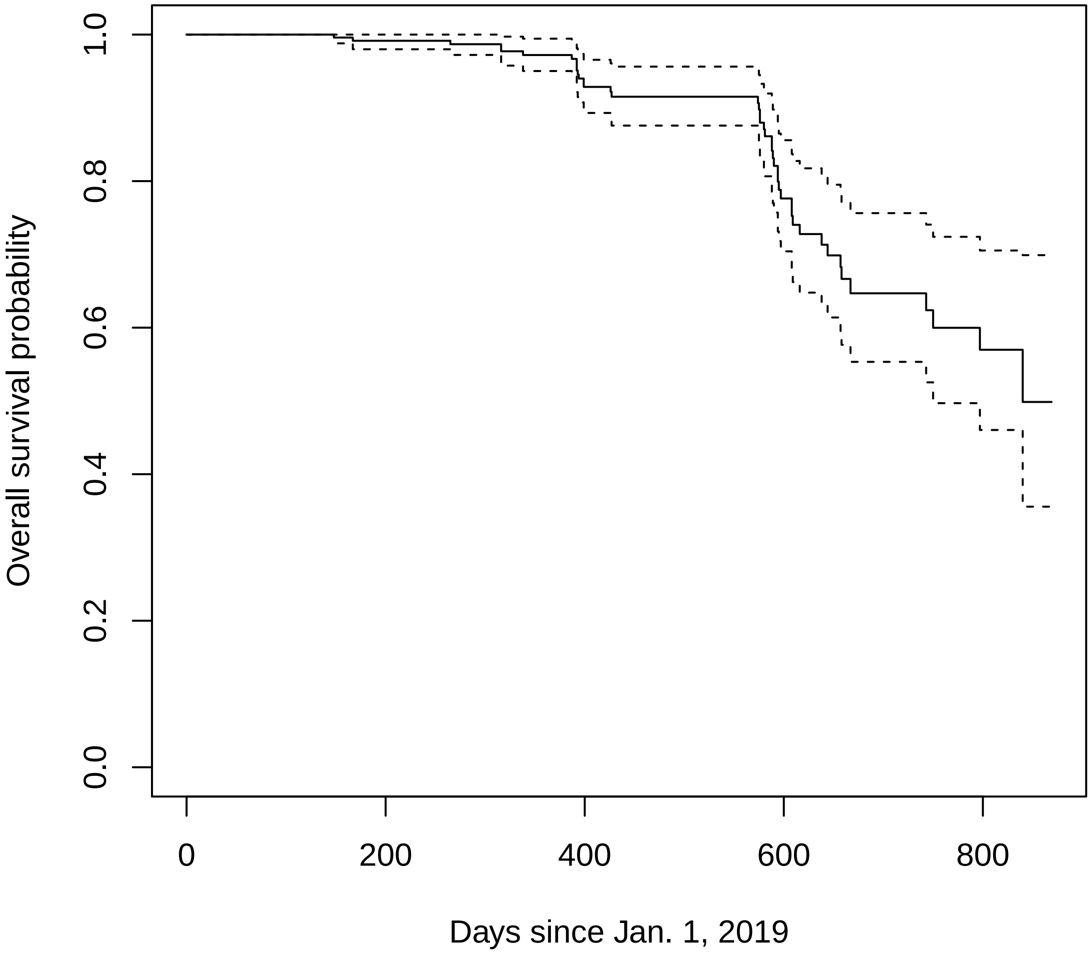

### Figure S3

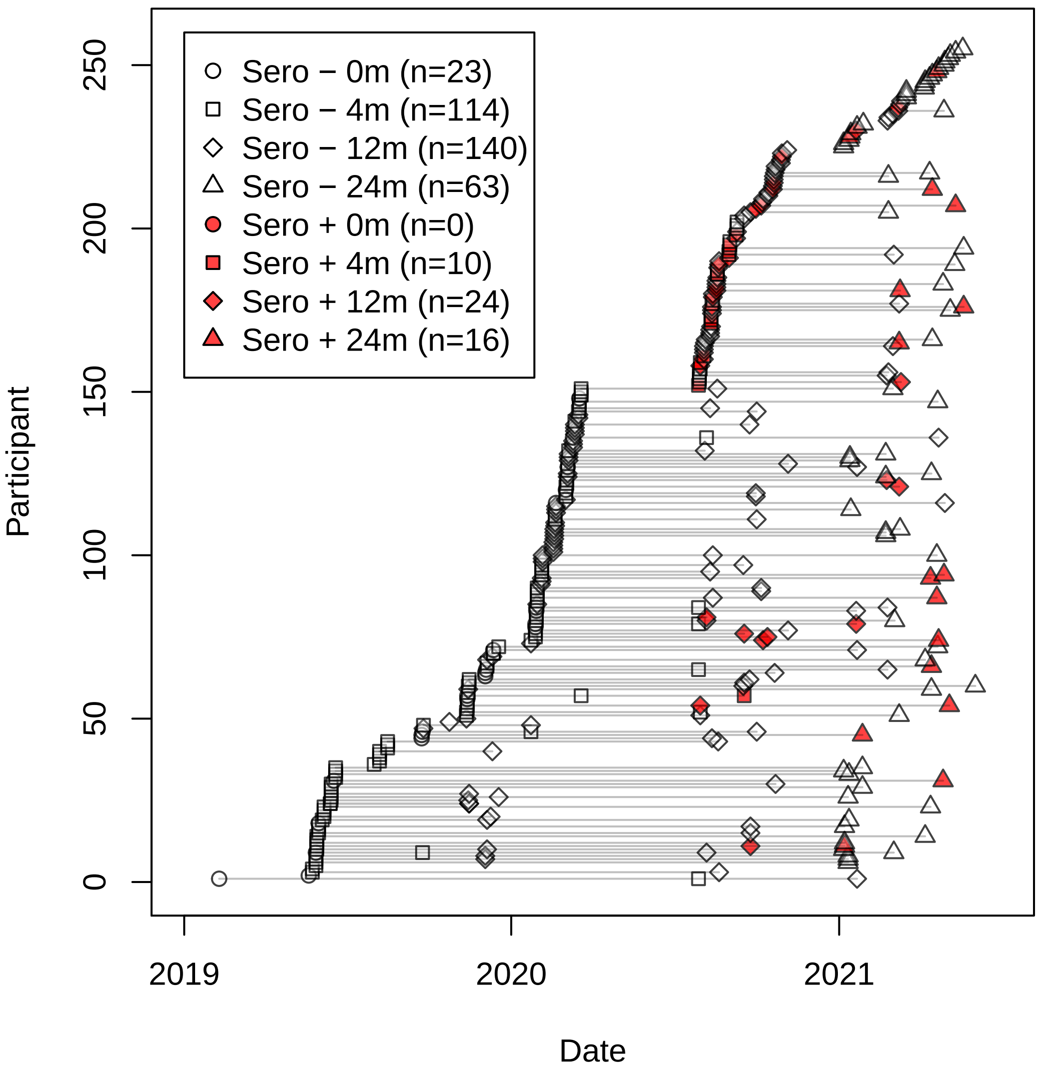
